# Food insecurity is associated with poor hypertension management in the Eastern Caribbean

**DOI:** 10.1101/2024.05.14.24307353

**Authors:** Carol R. Oladele, Neha Khandpur, Deron Galusha, Sanya Nair, Saria Hassan, Josefa Martinez-Brockman, Marcella Nunez-Smith, Rafael Perez-Escamilla

**Affiliations:** Equity Research and Innovation Center, Yale School of Medicine, New Haven, CT, USA; Wageningen University and Research, Wageningen, The Netherlands; Emory University School of Medicine, Emory Rollins School of Public Health, Atlanta, GA, USA; Department of Social and Behavioral Sciences, Yale School of Public Health, New Haven, CT, USA

## Abstract

**Background:** Limited evidence exists on the association between food insecurity (FI) and blood pressure control in the Caribbean despite the high burden of both. The objective of this study is to examine the relationship between FI and hypertension prevalence, awareness, and control in the Eastern Caribbean.

**Methods and Findings:** We conducted a cross-sectional analysis of baseline data (2013-2018) from the Eastern Caribbean Health Outcomes Research Network Cohort Study (n=2961). Food insecurity was measured using the Latin American and Caribbean Food Security Scale (ELCSA) and classified as 0=no FI, 1-6 mild/moderate FI, and 7-9 severe FI. Hypertension was defined by the Seventh Report of the Joint National Committee on Prevention. Logistic regression modeling was conducted to examine the relationship between FI and hypertension prevalence, awareness, and control, adjusting for age, sex, educational attainment, site, and usual source of care. Prevalence of FI was 28 percent among participants and was higher in Puerto Rico and Trinidad and Tobago compared to other sites. Seventeen percent of the participants experienced low, 6 percent moderate, and 4 percent experienced severe FI. Aggregate model results showed no association between FI and hypertension outcomes. Sex-stratified results showed moderate (OR=2.65, CI=1.25-5.65) and severe FI (OR=3.69, CI=1.20-11.31) were positively associated with lack of control among women.

Limitations of this study include the cross-sectional design, small sample size, and the average age of our cohort. Cross-sectional design precluded the ability to make inferences about temporality between FI and HTN prevalence and awareness. Small sample size precluded the ability to detect statistically significant differences despite strong odds ratios for model results like lack of control.

**Conclusions:** Findings align with prior evidence of greater FI prevalence among women and negative health impact. Nutrition policies are needed to reduce the overall FI burden in the Caribbean and increase access to affordable, nutritious foods.

## INTRODUCTION

Food insecurity (FI), defined by the WHO as an economic or social condition of limited or uncertain access to adequate food, is an increasing problem worldwide. Estimates from the Food and Agriculture Organization of the United Nations show that 2.3 billion people (29%) experienced moderate or severe food insecurity in 2021.^1^ Evidence also shows a widening gender disparity.^1^ FI has increased during the last ten years, with much of the increase stemming from the past three years during the COVID-19 pandemic. An estimated 41 percent of households experienced food insecurity in 2020, an increase from 32 percent pre-pandemic.^2^ There has also been a 38 percent increase in the number of people who reported going without food for an entire day between 2021 and 2022.^3^ Although it is well known that FI is associated with adverse health outcomes, including hypertension, a leading cause of morbidity in the region,^4^ there is limited evidence on the health impact of FI in hypertension management in the Caribbean region despite it being strongly affected by both FI and hypertension.

Several factors contribute to FI in the Caribbean, including a strong reliance on imported food,^5^ poverty,^5^ and climate change impacts.^5^ Countries in this region import 15 to 95 percent of their food. The high reliance on imported food poses FI challenges as these foods tend to be expensive, of limited nutritional value, and oftentimes ultra-processed. Our prior work in the Eastern Caribbean showed that affordability was a main determinant of fruit and vegetable consumption, which primarily affected individuals who experienced FI.^6^ Increasing poverty due to weak economic markets and joblessness has also been correlated with higher rates of FI.^7^ Extreme weather events due to climate change has affected food production and availability in the region, which in turn, has led to increased reliance on food imports.^5^ These interrelated factors have wide-ranging consequences, including poor dietary quality, a primary risk factor for diet-sensitive diseases like hypertension, and the inability of people to manage them properly.^8^ Given the prevalence of FI, evidence of negative health consequences, and paucity of evidence in the region, we sought to examine the relationship between FI and the hypertension cascade. Specifically, we aimed to understand the association between FI and prevalence, awareness, and control of hypertension.

## METHODS

### Data source and study sample

We analyzed baseline data from the Eastern Caribbean Health Outcomes Research Network Cohort Study (ECS) for this study. The ECS is an ongoing longitudinal cohort study conducted across four Caribbean sites that aims to identify novel risk and protective factors for non-communicable diseases in the Eastern Caribbean region. The sites included are the U.S. Virgin Islands, Puerto Rico, Trinidad and Tobago, and Barbados. The cohort was empaneled between 2013 and 2018 across the four sites using varied methods to obtain randomized samples in each site (n=2,961)^9^. Eligible participants were English or Spanish-speaking community-dwelling adults 40 years of age and older who had been residents of the island for at least 10 years and intended to live on the island for the next 5 years. ECS participants were included in the current study if they had complete blood pressure and food insecurity data at baseline.

### Measures

Our main exposure was food insecurity. Food insecurity was measured using the 9-item version of the Latin American and Caribbean Food Security Scale (ELCSA) previously validated in the Caribbean.^10^ The ELCSA captured household food insecurity within the past 90 days. Response options were binary (yes/no), and one point was given for each question with a “yes” response. Responses were summed for each participant and ranged from 0 to 9. Those who scored 0 were classified as having no food insecurity, 1-6 as having mild/moderate food insecurity, and 7-9 as having severe food insecurity. The ninth ELCSA item addresses the social acceptability dimension of FI and was included as part of ELCSA’s summative score based on psychometric testing included. Rasch’s modeling of the ELCSA scale in the ECS sample indicated that the full 9-item scale was the best fit^11^. The Cronbach’s Alpha for the scale was 0.90.

Our main outcome was hypertension. Hypertension was defined using guidelines established by the Seventh Report of the Joint National Committee^12^ and Caribbean Health Research Council^13^, which were clinical guidelines used to identify hypertension during the ECS data collection period. Hypertension was assessed using self-report and clinical assessment data. Details about the clinical assessment processes have been previously described.^14^ Participants were classified as having hypertension if they had systolic blood pressure ≥ 140 mmHg or diastolic blood pressure ≥ 90 mmHg during the clinical exam or answered “yes” to the following question, “Has a doctor or other health provider EVER told you that you have high blood pressure?” and reported taking blood pressure lowering medication. Participants who responded “no” to this question and had blood pressure <140 mmHg and diastolic blood pressure < 90 mmHg were classified as not having hypertension.

Hypertension awareness was assessed as answering “yes” to the question, “Has a doctor or other health provider EVER told you that you have high blood pressure?” and reported taking blood pressure lowering medication. Responses of “yes” were categorized as “Aware.” Participants who responded “no” and had elevated systolic or diastolic blood pressure during the clinical exam were categorized as “unaware.” Hypertension control was determined among those who were aware and reported taking medication. Control was assessed using blood pressure values from the clinical exam and defined as having systolic blood pressure <140 mmHg and diastolic blood pressure <90 mmHg.

Covariates examined included demographic characteristics, healthcare utilization, and site. Demographic characteristics included age, sex, and educational attainment, which were measured via self-report during the ECS baseline survey. We categorized age into four groups (40-49, 50-59, 60-69, and 70+). Sex was measured on the baseline survey using the following question, “What sex were you at birth?” Educational attainment was measured using the question, “What is the highest year of school that you completed?” Responses were categorized into less than high school (or secondary school), high school graduate, some college, and college and higher. Usual source of care was used to characterize healthcare utilization and was measured using the question, “Is there one place you usually go when you need routine or non-emergent/non-emergency care (for example, regular check-up)”? Responses were categorized into three categories: none, one, or more.

### Statistical Analysis

Univariate analyses were conducted to examine the frequency distribution of variables. We conducted bivariate analyses using chi-square tests to examine the distribution of FI across demographic and healthcare characteristics. Unadjusted and adjusted logistic regression analyses were performed to determine associations between FI and hypertension prevalence, awareness, and control. Adjusted models included age, sex, educational attainment, site, and usual source of care. These covariates were selected based on evidence from prior literature.^15–17^ Sex-stratified modeling was performed to determine potential differences in the impact of FI for men and women. Analyses were conducted using SAS statistical software, version 9.4 (Research Triangle Institute, Research Triangle Park).

### Ethics Statement

The ECS study was approved by the Yale University Human Subjects Investigation Committee (protocol 2000026077), the Institutional Review Boards of the University of Puerto Rico Medical Sciences Campus (protocol 2290033151R001), the University of the Virgin Islands (protocol 460911-18), and the University of the West Indies Cave Hill Campus (IRB Number: 171102-A), as well as by the Ministry of Health of Trinidad and Tobago. Formal written consent was obtained from study participants. The current analysis was approved by the Data Access and Scientific Review Committee of the ECS. This study was reported according to STROBE guidelines.

## RESULTS

Our final analytic dataset included 2,323 observations after excluding 638 participants who had missing FI and blood pressure data (Fig 1). The mean age of participants was 57, 65 percent were women, 35 percent had less than a high school education, and most (85%) had one or more usual places they received care. The overall prevalence of FI was 28 percent among participants, and it was higher in Puerto Rico and Trinidad and Tobago compared to other sites. Among those with FI, 17 percent experienced low, 6 percent moderate, and 4 percent experienced severe FI. Bivariate analyses showed statistically significant differences in FI by age, sex, educational attainment, and site (Table 1). Those living in households with moderate or severe FI were younger compared to those who were living in households that were food secure or had low FI. Women and those with less than a high school education were more likely to experience low, moderate, or severe FI compared to men and those with higher educational attainment (p<0.05). Table 2 presents prevalence estimates and logistic regression model results for the relationship between FI and hypertension prevalence, awareness, and control. Hypertension prevalence was 54, 51, and 52 percent among persons with low, moderate, and severe FI, respectively.

**Figure 1:**
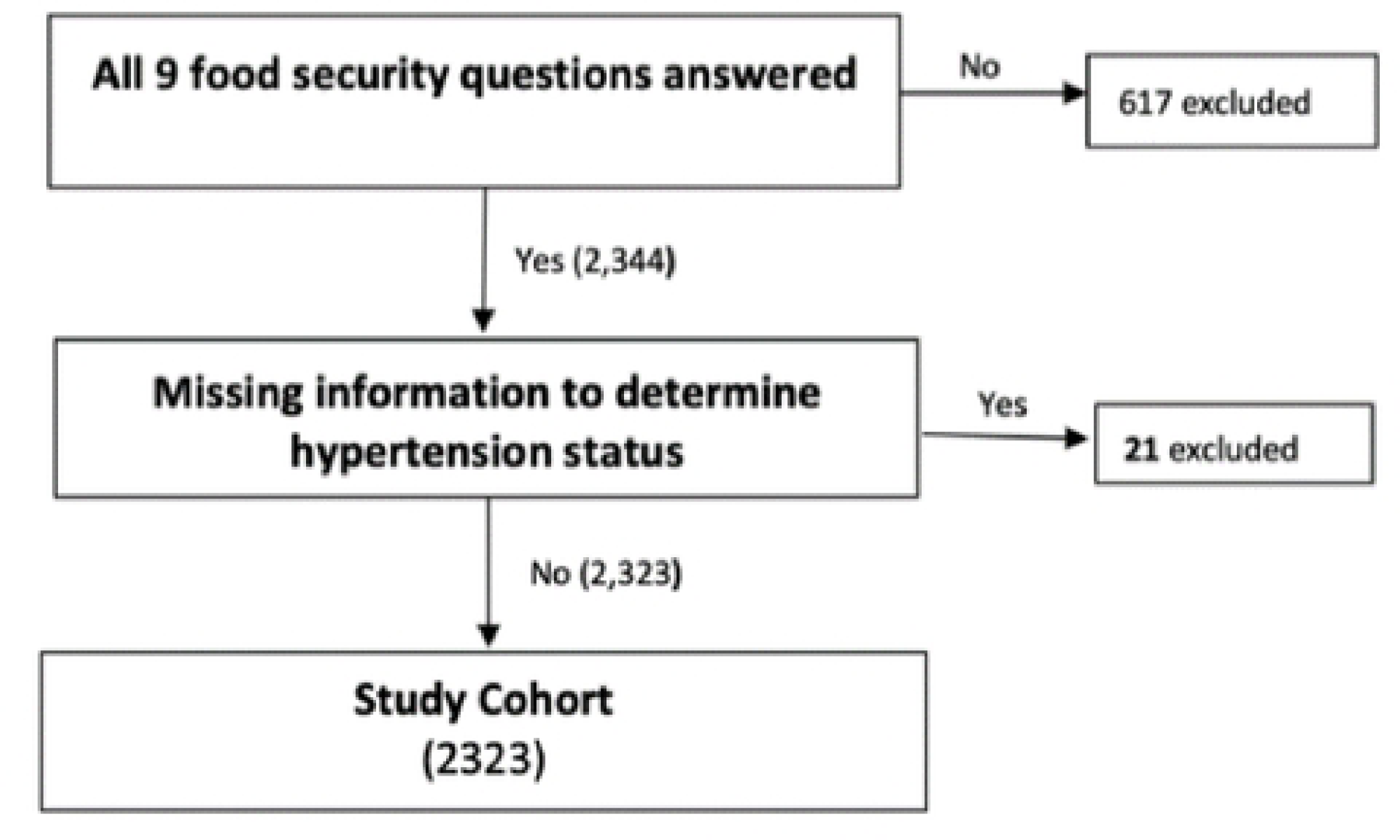
Construction of Study Cohort.

**Table 1.** ECS participant characteristics by food security status.

| Characteristics | Total | Secure | Low | Moderate | Severe | p-value |
| --- | --- | --- | --- | --- | --- | --- |
|  | 2323 | 1683<br>(72.4%) | 394<br>(17.0%) | 144<br>(6.2%) | 102<br>(4.4%) |  |
| Mean age (SD) | 57.3 (10.5) | 58.8 (10.6) | 54.1 (9.5) | 51.6 (8.0) | 52.9 (7.4) | <0.0001 |
| Sex |  |  |  |  |  | 0.0279 |
| Men | 817 (35.2) | 614 (36.5) | 133 (33.8) | 35 (24.3) | 35 (34.3) |  |
| Women | 1506 (64.8) | 1069 (63.5) | 261 (66.2) | 109 (75.7) | 67 (65.7) |  |
| Educational attainment |  |  |  |  |  | <.0001 |
| < HS | 789 (34.8) | 541 (32.9) | 151 (39.7) | 55 (39) | 42 (42.0) |  |
| HS Grad | 540 (23.8) | 395 (24.0) | 88 (23.2) | 32 (22.7) | 25 (25.0) |  |
| Some College | 501 (22.1) | 348 (21.2) | 88 (23.2) | 38 (27) | 27 (27.0) |  |
| College+ | 436 (19.2) | 361 (21.9) | 53 (13.9) | 16 (11.4) | 6 (6.0) |  |
| Site |  |  |  |  |  | <.0001 |
| Barbados | 703 (30.3) | 552 (32.8) | 92 (23.4) | 36 (25) | 23 (22.6) |  |
| Puerto Rico | 735 (31.6) | 566 (33.6) | 98 (24.9) | 41 (28.5) | 30 (29.4) |  |
| Trinidad and Tobago | 716 (30.8) | 452 (26.9) | 166 (42.1) | 55 (38.2) | 43 (42.2) |  |
| US Virgin Islands | 169 (7.3) | 113 (6.7) | 38 (9.6) | 12 (8.3) | 6 (5.9) |  |
| Usual source of healthcare |  |  |  |  |  | 0.5157 |
| None | 339 (14.7) | 235 (14.0) | 61 (15.5) | 25 (17.4) | 18 (17.7) |  |
| One or more | 1974 (85.3) | 1438 (86.0) | 333 (84.5) | 119 (82.6) | 84 (82.4) | .. |

**Table 2.** Association between level of food insecurity and hypertension outcomes.

|  | Level of Food Insecurity |  |  |  |
| --- | --- | --- | --- | --- |
|  | Secure (N=1683) | Low (N=394) | Moderate (N=144) | Severe (N=102) |
| Prevalence of Hypertension | 991/1683 (58.9%) | 211/394 (53.6%) | 73/144 (50.7%) | 53/102 (52%) |
| Unadjusted OR (95% CI) | 1.00 | 0.81 (0.65-1) | 0.72 (0.51-1.01) | 0.76 (0.51-1.13) |
| Adjusted* OR (95% CI) | 1.00 | 1.08 (0.85-1.38) | 1.1 (0.76-1.59) | 1.01 (0.66-1.54) |
| Lack of Awareness of Hypertension | 346/991 (34.9%) | 82/211 (38.9%) | 23/73 (31.5%) | 19/53 (35.8%) |
| Unadjusted OR (95% CI) | 1.00 | 1.19 (0.87-1.61) | 0.86 (0.52-1.43) | 1.04 (0.59-1.85) |
| Adjusted* OR (95% CI) | 1.00 | 0.84 (0.59-1.19) | 0.68 (0.38-1.21) | 0.68 (0.36-1.3) |
| Lack of Control of Hypertension | 365/645 (56.6%) | 73/129 (56.6%) | 34/50 (68%) | 25/34 (73.5%) |
| Unadjusted OR (95% CI) | 1.00 | 1 (0.68-1.46) | 1.63 (0.88-3.01) | 2.13 (0.98-4.63) |
| Adjusted* OR (95% CI) | 1.00 | 0.98 (0.65-1.48) | 2.18 (1.13-4.21) | 2.31 (0.99-5.37) |

\* Adjusted for age, sex, education level, site, and usual source of care

Unadjusted and adjusted model results for FI and hypertension prevalence were not statistically significant.

Results for lack of awareness showed that 35 percent of those who lived in food-secure households and 39 percent of low food-insecure households were unaware of their hypertension, compared to 32 percent and 36 percent among those with moderate and severe FI. Unadjusted and adjusted model results showed that lack of hypertension awareness was not significantly associated with FI.

Results for lack of control showed that 57 percent of households with low FI, 68 percent of those with moderate FI, and 73 percent of those with severe FI did not have their hypertension controlled, compared to 57 percent among persons who were food secure. Adjusted model results for control showed moderate FI was associated with higher odds (OR=2.18; CI=1.13-4.21) of poorly controlled hypertension compared to those who were food secure.

Table 3 presents sex-stratified model results. Results showed that overall, women who experienced moderate or severe FI were more likely to have hypertension and poor control compared to men. Fifty-four and 58 percent of women with moderate and severe FI had hypertension, compared to 40 percent among men with moderate and severe FI. Similarly, results for lack of control showed that women with moderate (72% vs. 43%) and severe FI (81% vs. 50%) were more likely to lack control of their hypertension compared to men. Unadjusted and adjusted model results showed statistically significant positive associations between moderate and severe FI and lack of control for women. Results showed that women with moderate and severe FI had 2.6- and 3.7 times greater odds of uncontrolled hypertension. Results for men were not statistically significant.

**Table 3.**
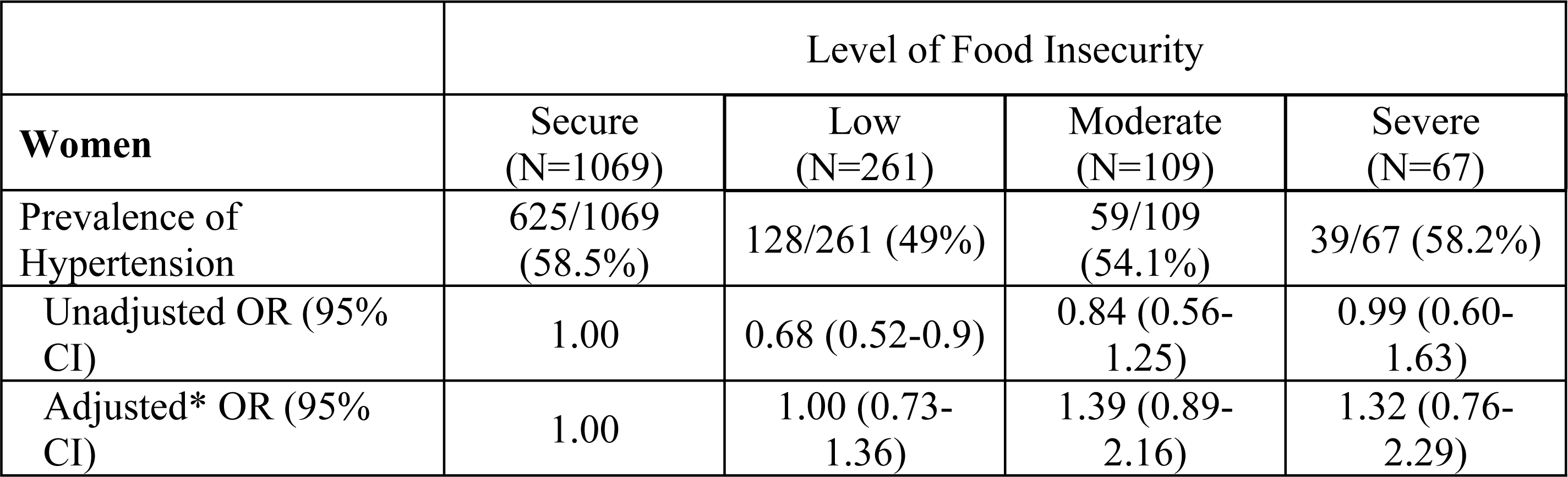
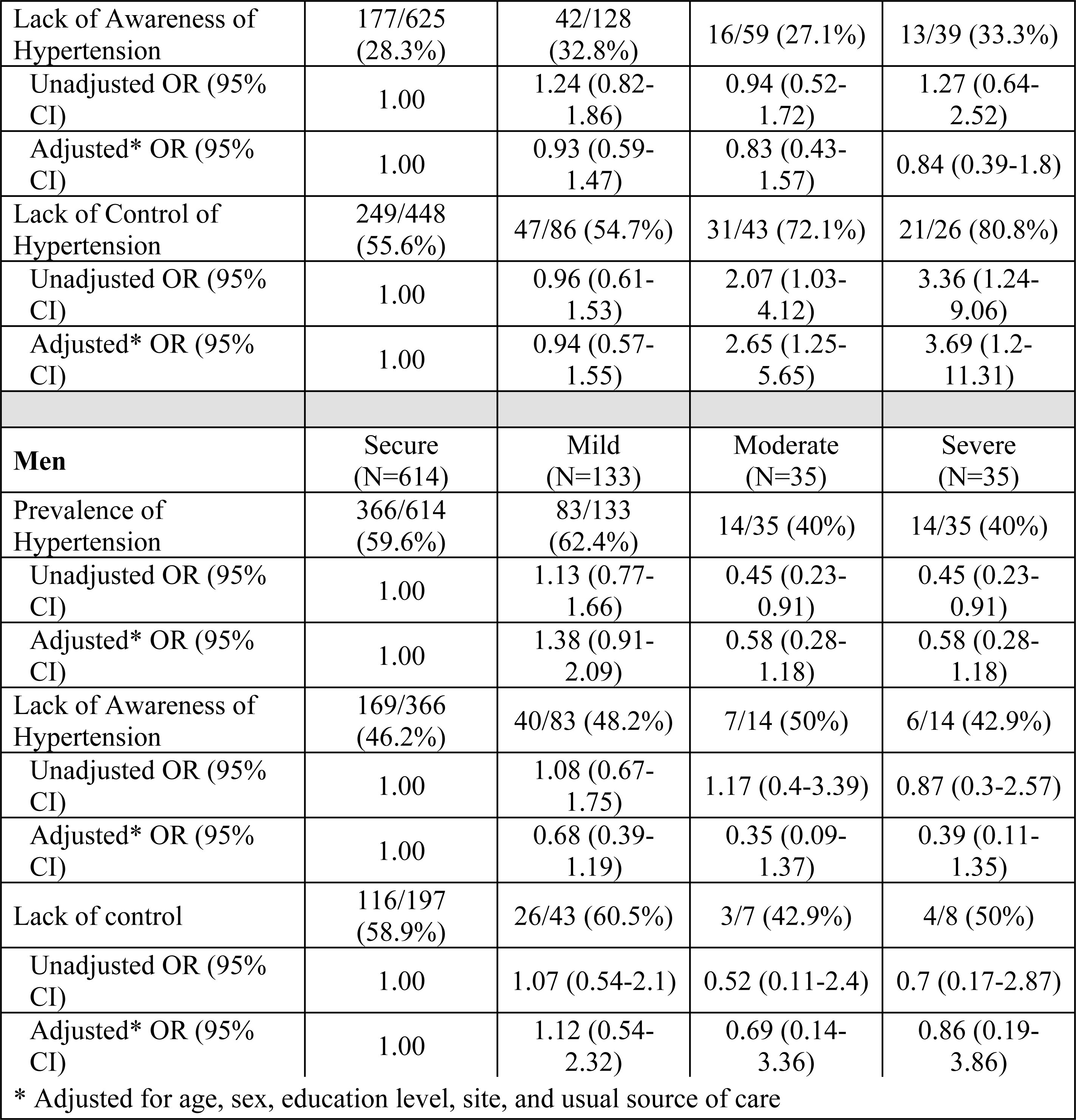
Gender-stratified models for food insecurity and hypertension control.

|  | Level of Food Insecurity |  |  |  |
| --- | --- | --- | --- | --- |
| **Women** | Secure<br>(N=1069) | Low<br>(N=261) | Moderate<br>(N=109) | Severe<br>(N=67) |
| Prevalence of Hypertension | 625/1069<br>(58.5%) | 128/261 (49%) | 59/109<br>(54.1%) | 39/67 (58.2%) |
| Unadjusted OR (95% CI) | 1.00 | 0.68 (0.52-0.9) | 0.84 (0.56-1.25) | 0.99 (0.60-1.63) |
| Adjusted* OR (95% CI) | 1.00 | 1.00 (0.73-1.36) | 1.39 (0.89-2.16) | 1.32 (0.76-2.29) |
| Lack of Awareness of Hypertension | 177/625 (28.3%) | 42/128 (32.8%) | 16/59 (27.1%) | 13/39 (33.3%) |
| Unadjusted OR (95% CI) | 1.00 | 1.24 (0.82-1.86) | 0.94 (0.52-1.72) | 1.27 (0.64-2.52) |
| Adjusted* OR (95% CI) | 1.00 | 0.93 (0.59-1.47) | 0.83 (0.43-1.57) | 0.84 (0.39-1.8) |
| Lack of Control of Hypertension | 249/448 (55.6%) | 47/86 (54.7%) | 31/43 (72.1%) | 21/26 (80.8%) |
| Unadjusted OR (95% CI) | 1.00 | 0.96 (0.61-1.53) | 2.07 (1.03-4.12) | 3.36 (1.24-9.06) |
| Adjusted* OR (95% CI) | 1.00 | 0.94 (0.57-1.55) | 2.65 (1.25-5.65) | 3.69 (1.2-11.31) |
| **Men** | Secure (N=614) | Mild (N=133) | Moderate (N=35) | Severe (N=35) |
| Prevalence of Hypertension | 366/614 (59.6%) | 83/133 (62.4%) | 14/35 (40%) | 14/35 (40%) |
| Unadjusted OR (95% CI) | 1.00 | 1.13 (0.77-1.66) | 0.45 (0.23-0.91) | 0.45 (0.23-0.91) |
| Adjusted* OR (95% CI) | 1.00 | 1.38 (0.91-2.09) | 0.58 (0.28-1.18) | 0.58 (0.28-1.18) |
| Lack of Awareness of Hypertension | 169/366 (46.2%) | 40/83 (48.2%) | 7/14 (50%) | 6/14 (42.9%) |
| Unadjusted OR (95% CI) | 1.00 | 1.08 (0.67-1.75) | 1.17 (0.4-3.39) | 0.87 (0.3-2.57) |
| Adjusted* OR (95% CI) | 1.00 | 0.68 (0.39-1.19) | 0.35 (0.09-1.37) | 0.39 (0.11-1.35) |
| Lack of control | 116/197 (58.9%) | 26/43 (60.5%) | 3/7 (42.9%) | 4/8 (50%) |
| Unadjusted OR (95% CI) | 1.00 | 1.07 (0.54-2.1) | 0.52 (0.11-2.4) | 0.7 (0.17-2.87) |
| Adjusted* OR (95% CI) | 1.00 | 1.12 (0.54-2.32) | 0.69 (0.14-3.36) | 0.86 (0.19-3.86) |
| * Adjusted for age, sex, education level, site, and usual source of care |  |  |  |  |

## DISCUSSION

This study aimed to determine the relationship between FI and hypertension prevalence, awareness, and control in the Eastern Caribbean. Our findings showed that FI prevalence was 28 percent and 10 percent of households experienced moderate or severe FI. Findings also showed sex differences in experiences with FI and the relationship between FI and hypertension control. Women were more likely to experience food insecurity, and a higher proportion of women with FI had hypertension compared to men. Findings also showed a statistically significant and positive association between FI and lack of hypertension control among women. These findings further substantiate FI as a significant public health issue in the Caribbean and demonstrate its role in poor blood pressure control. In addition, our findings highlight the differential impact of FI on health outcomes among women. This study contributes to a growing evidence base on the health impacts of FI in the Caribbean, where evidence is limited yet needed to guide the creation of region-specific solutions.

Our finding that showed a relationship between FI and poor blood pressure control is novel within the FI and chronic disease literature. While prior studies have examined associations between FI and chronic disease management, the preponderance of evidence is focused on diabetes prevalence and management.^18,19^ Fewer studies have examined the association between FI and hypertension or examined the role of FI in poor blood pressure management, though it has been linked to poor dietary quality.^20,21^ Findings from studies conducted in the US and other high-income countries show mixed findings for the relationship between FI and hypertension prevalence, which may be attributed to variation in how blood pressure is measured (self-report vs. clinical measurement) in studies. Most fail to demonstrate a statistically significant association. Evidence on the relationship between FI and blood pressure management is scant. Studies that have examined this association suggest a positive association.^20–23^ Potential mechanistic pathways between FI and poor blood pressure include poor dietary quality, inflammation, and gut microbiome disruptions.^24–26^

Findings on sex differences in FI prevalence and the differential impact of FI on blood pressure control among women are consistent with prior studies. The consistency of these findings spans context, culture, race, and ethnicity. Several explanations have been offered. Gendered roles and inequities in earning power and household responsibilities are some explanations for the observed differences in FI between men and women.^27^ In most cases, women are responsible for food purchasing and preparation. Some evidence suggests that women are more likely to sacrifice in households experiencing FI by employing strategies like reducing food consumption, reducing the diversity of foods they consume, and consuming cheaper energy-dense foods to provide for others in the household.^27^ Evidence also suggests women are less likely to absorb economic and household shocks related to FI due to inequities in access to assets and resources to cope with FI.28 Mounting evidence that shows sex differences in the relationship between FI and obesity^29–31^ a risk factor for hypertension, may offer another explanation. Evidence shows a positive relationship among women^32,33^ but not men, which may complicate hypertension management among women with FI.^34^

The current findings are generalizable to populations represented in the ECHORN sites and have several implications. The high burden of FI across the region and the demonstrated association with poor blood pressure control suggests an urgent need for policy and healthcare system interventions. A recent review of nutrition interventions and policies in the small island developing states, including those in the Caribbean, highlighted the paucity of nutrition policies in the region and the overall low proportion of states where policies were fully implemented.^35^

### Limitations

This study has few limitations. The cross-sectional design of our study precludes the ability to make inferences about temporality in the relationship between FI and HTN. This may have contributed to our inability to observe a relationship between FI and HTN prevalence and awareness. The small sample size available to examine associations between moderate and severe FI and hypertension outcomes may have precluded the ability to detect statistically significant differences despite observing strong odds ratios for some model results like lack of control. Given the age of our cohort and findings that show higher FI among younger persons, our study may underestimate the overall true burden of FI in the Caribbean region. Despite these limitations, findings remain salient to efforts to reduce FI and improve blood pressure management in the region. Future studies will seek to identify food environment and individual factors that contribute to poor blood pressure management among individuals experiencing FI.

### Public Health Implications

Findings have implications for healthcare system solutions to identify and address FI and other complex social factors that contribute to poor blood pressure control. In addition, an examination of clinical factors that contribute to a lack of blood pressure control is warranted.

## Data Availability

The corresponding author had full access to all the data in the study and takes responsibility for its integrity and the data analysis. Data described in the manuscript will be made available upon reasonable request.

https://www.echorn.org/request-echorn-data

## ACKNOWLEDGMENTS

None.

## Disclosure of Which Tasks Each Author Completed

CO, NK, RPE, SH, and JM conceptualized the study. MNS contributed to data acquisition. CO, RPE, and DG analyzed data. CO, NK, SH, SN, and JM interpreted the results. CO contributed to drafting and writing. NK, MNS, SH, SN, and JM contributed to the critical review. MNS and RPE approved the final draft of the paper. All authors read and approved the final manuscript.

## Conflicts of Interest

The study sponsor has no role in the study design, collection, analysis, and interpretation of the data. To the best of our knowledge, there are no relevant conflicts of interest, financial or otherwise, relevant to this study and the publication thereof to declare.

## Notes

### Competing Interest Statement

The authors have declared no competing interest.

### Clinical Trial

NA

### Author Declarations

The ECS study was approved by the Yale University Human Subjects Investigation Committee, the Institutional Review Boards of the University of Puerto Rico Medical Sciences Campus, the University of the Virgin Islands, and the University of the West Indies Cave Hill Campus, as well as by the Ministry of Health of Trinidad and Tobago. The current analysis was approved by the Data Access and Scientific Review Committee of the ECS. This study was reported according to STROBE guidelines.

## REFERENCES

1. UN Report: Global hunger numbers rose to as many as 828 million in 2021. https://www.fao.org/newsroom/detail/un-report-global-hunger-SOFI-2022-FAO/en

2. World Food Programme. Sharp rise in food insecurity in the Caribbean, survey finds. Accessed January 20, 2023. https://www.wfp.org/news/sharp-rise-food-insecurity-caribbean-survey-finds

3. Programme WF. Caribbean Food Security and Livelihoods Impact Survey. 2023.

4. Oladele CR, Martinez J, Nunez-Smith M. Abstract P094: Food Insecurity And Elevated Blood Pressure In The Eastern Caribbean. Circulation. 2021;143(Suppl_1):AP0941-AP094. doi:doi:10.1161/circ.143.suppl_1.P094

5. Mohammadi E, Singh SJ, McCordic C, Pittman J. Food Security Challenges and Options in the Caribbean: Insights from a Scoping Review. Anthropocene Science. 2022/03/01 2022;1(1):91-108. doi:10.1007/s44177-021-00008-8

6. Oladele CR, Colón-Ramos U, Galusha D, et al. Perceptions of the local food environment and fruit and vegetable intake in the Eastern Caribbean Health Outcomes research Network (ECHORN) Cohort study. Prev Med Rep. Apr 2022;26:101694. doi:10.1016/j.pmedr.2022.101694

7. Food Insecurity in the Caribbean continues on upward trajectory, CARICOM-WFP survey finds. World Food Programme. https://www.wfp.org/news/food-insecurity-caribbean-continues-upward-trajectory-caricom-wfp-survey-finds

8. Grilo SA, Shallcross AJ, Ogedegbe G, Odedosu T, Levy N, Lehrer S. Food Insecurity and Effectiveness of Behavioral Interventions to Reduce Blood Pressure, New York City, 2012-2013. Preventing Chronic Disease. 2015;12:E16. doi:10.5888/pcd12.140368

9. Thompson T, Maharaj R, Nunez M, Nazario C, Adams O, Nunez-Smith M. Non-communicable diseases III: the Eastern Caribbean health outcomes research network (ECHORN) cohort study. The West Indian medical journal. 2018;67:40.

10. Perez-Escamilla R, Dessalines M, Finnigan M, Pachon H, Hromi-Fiedler A, Gupta N. Household food insecurity is associated with childhood malaria in rural Haiti. The Journal of nutrition. Nov 2009;139(11):2132–8. doi:10.3945/jn.109.108852

11. Martinez-Brockman J, Hromi-Fiedler A, Galusha D, et al. Risk Factors for Household Food Insecurity in the Eastern Caribbean Health Outcomes Research Network (ECHORN) Cohort Study. Frontiers Public Health. 2023.

12. Chobanian AV, Bakris GL, Black HR, Cushman WC, Green LA, Izzo JL. The seventh report of the Joint National Committee on Prevention, detection, evaluation, and treatment of high blood pressure: the JNC 7 report. JAMA: the journal of the American Medical Association. 2003;289doi:10.1001/jama.289.19.2560

13. council CCMR. Managing Hypertension in Primary Care in the Caribbean. 1998.

14. Spatz ES, Martinez-Brockman JL, Tessier-Sherman B, et al. Phenotypes of Hypertensive Ambulatory Blood Pressure Patterns: Design and Rationale of the ECHORN Hypertension Study. Ethn Dis. Fall 2019;29(4):535–544. doi:10.18865/ed.29.4.535

15. Connelly PJ, Delles C. Journal of Human Hypertension special issue on sex and gender differences in hypertension. Journal of Human Hypertension. 2023/08/01 2023;37(8):587-588. doi:10.1038/s41371-023-00847-5

16. Lopez-Lopez JP, Cohen DD, Alarcon-Ariza N, et al. Ethnic Differences in the Prevalence of Hypertension in Colombia: Association With Education Level. Am J Hypertens. Jul 1 2022;35(7):610–618. doi:10.1093/ajh/hpac051

17. Mills KT, Stefanescu A, He J. The global epidemiology of hypertension. Nat Rev Nephrol. Apr 2020;16(4):223–237. doi:10.1038/s41581-019-0244-2

18. Thomas MK, Lammert LJ, Beverly EA. Food Insecurity and its Impact on Body Weight, Type 2 Diabetes, Cardiovascular Disease, and Mental Health. Curr Cardiovasc Risk Rep. 2021;15(9):15. doi:10.1007/s12170-021-00679-3

19. Vaccaro JA, Huffman FG. Sex and Race/Ethnic Disparities in Food Security and Chronic Diseases in U.S. Older Adults. Gerontol Geriatr Med. Jan-Dec 2017;3:2333721417718344. doi:10.1177/2333721417718344

20. Beltrán S, Pharel M, Montgomery CT, López-Hinojosa IJ, Arenas DJ, DeLisser HM. Food insecurity and hypertension: A systematic review and meta-analysis. PLoS One. 2020;15(11):e0241628. doi:10.1371/journal.pone.0241628

21. Pérez-Escamilla R, Villalpando S, Shamah-Levy T, Méndez-Gómez Humarán I. Household food insecurity, diabetes and hypertension among Mexican adults: results from Ensanut 2012. Salud Publica Mex. 2014;56 Suppl 1:s62–70. doi:10.21149/spm.v56s1.5167

22. Hojaji E, Aghajani M, Zavoshy R, Noroozi M, Jahanihashemi H, Ezzeddin N. Household food insecurity associations with pregnancy hypertension, diabetes mellitus and infant birth anthropometric measures: a cross-sectional study of Iranian mothers. Hypertens Pregnancy. May 2021;40(2):109–117. doi:10.1080/10641955.2021.1874010

23. Liu Y, Eicher-Miller HA. Food Insecurity and Cardiovascular Disease Risk. Curr Atheroscler Rep. Mar 27 2021;23(6):24. doi:10.1007/s11883-021-00923-6

24. Li J, Zhao F, Wang Y, et al. Gut microbiota dysbiosis contributes to the development of hypertension. Microbiome. 2017/02/01 2017;5(1):14. doi:10.1186/s40168-016-0222-x

25. Yan Q, Gu Y, Li X, et al. Alterations of the Gut Microbiome in Hypertension. Original Research. Frontiers in Cellular and Infection Microbiology. 2017-August-24 2017;7 doi:10.3389/fcimb.2017.00381

26. Juul F, Vaidean G, Parekh N. Ultra-processed Foods and Cardiovascular Diseases: Potential Mechanisms of Action. Adv Nutr. Oct 1 2021;12(5):1673–1680. doi:10.1093/advances/nmab049

27. Broussard NH. What explains gender differences in food insecurity? Food Policy. 2019/02/01/ 2019;83:180-194. 10.1016/j.foodpol.2019.01.003

28. Seligman HK, Laraia BA, Kushel MB. Food insecurity is associated with chronic disease among low-income NHANES participants. The Journal of nutrition. Feb 2010;140(2):304–10. doi:10.3945/jn.109.112573

29. Hernandez DC, Reesor L, Murillo R. Gender Disparities in the Food Insecurity-Overweight and Food Insecurity-Obesity Paradox among Low-Income Older Adults. J Acad Nutr Diet. Jul 2017;117(7):1087–1096. doi:10.1016/j.jand.2017.01.014

30. Hernandez DC, Reesor LM, Murillo R. Food insecurity and adult overweight/obesity: Gender and race/ethnic disparities. Appetite. Oct 1 2017;117:373–378. doi:10.1016/j.appet.2017.07.010

31. Koller EC, Egede LE, Garacci E, Williams JS. Gender Differences in the Relationship Between Food Insecurity and Body Mass Index Among Adults in the USA. J Gen Intern Med. Dec 2022;37(16):4202–4208. doi:10.1007/s11606-022-07714-y

32. Monroy Torres R, Castillo-Chávez AM, Ruíz-González S. [Food insecurity and its association with obesity and cardiometabolic risks in Mexican women]. Nutr Hosp. Apr 19 2021;38(2):388–395. Inseguridad alimentaria y su asociación con la obesidad y los riesgos cardiometabólicos en mujeres mexicanas. doi:10.20960/nh.03389

33. Schlüssel MM, Silva AA, Pérez-Escamilla R, Kac G. Household food insecurity and excess weight/obesity among Brazilian women and children: a life-course approach. Cad Saude Publica. Feb 2013;29(2):219–26. doi:10.1590/s0102-311x2013000200003

34. Gooding HC, Walls CE, Richmond TK. Food insecurity and increased BMI in young adult women. Obesity (Silver Spring). Sep 2012;20(9):1896–901. doi:10.1038/oby.2011.233

35. Brown CR, Rocke K, Murphy MM, Hambleton IR. Interventions and policies aimed at improving nutrition in Small Island Developing States: a rapid review. Rev Panam Salud Publica. 2022;46:e33. doi:10.26633/rpsp.2022.33

